# A35R and H3L are Serological and B Cell Markers for Monkeypox Infection

**DOI:** 10.1101/2022.08.22.22278946

**Authors:** Ron Yefet, Nadav Friedel, Hadas Tamir, Ksenia Polonsky, Michael Mor, David Hagin, Eli Sprecher, Tomer Israely, Natalia T Freund

**Affiliations:** Department of Microbiology and Clinical Immunology, Faculty of Medicine, Tel Aviv University, Tel Aviv, Israel; Division of Dermatology, Tel-Aviv Sourasky Medical Center, Tel Aviv, Israel; Department of Infectious Diseases, The Israel Institute for Biological Research, Ness Ziona, Israel; Edmond J. Safra Center for Bioinformatics at Tel-Aviv University, Tel Aviv, Israel; Department of Immunology, Tel Aviv Sourasky Medical Center, Tel Aviv, Israel; Department of Human Molecular Genetics and Biochemistry, Faculty of Medicine, Tel-Aviv University, Tel Aviv, Israel

## Abstract

Monkeypox virus (MPXV) resides in two forms, mature and enveloped virions, and depending on it, distinct proteins are displayed on the viral surface. We expressed in mammalian cells two MPXV antigens from the mature form and two MPXV antigens from the enveloped form and tested their reactivity to sera of 11 MPXV convalescent donors diagnosed in Israel during May-June 2022 and collected 33-62 days post infection. While only 4 out of 11 donors neutralized the related Vaccinia Lister strain, all MPXV recoverees demonstrated a strong serological response to a 124-amino acid truncation of the A35R antigen, and to a 276-amino acid truncation of the H3L antigen. Moreover, A35R- and H3L-specific B cells ranging from 0.03-0.5% and 0.01-0.35% of IgG+CD19+ cells, respectively, were detected in all 11 MPXV donors (A35R), and in 8 out of 11 donors (H3L). Therefore, A35R and H3L represent MPXV immune targets, and can be used in a simple heat-inactivated serological enzyme-linked immunosorbent assay for the identification of recent MPXV infection.

## Main Text

Monkeypox virus (MPXV) is a member of the Orthopoxvirus genus ^1,2^ and is responsible for Monkeypox disease, including the current Monkeypox outbreak, which is the largest recorded outbreak in non-endemic countries to date ^3^. As of August 15^th^ 2022, 35,514 people outside Africa were infected, with most cases diagnosed in Spain and the US ^4^. The infection results in blisters, fever and discomfort, with case mortality rate of less than 0.5% (for the current outbreak). Diagnosis of MPXV infection is based on polymerase chain reaction (PCR) to detect the MPXV nucleic acids ^5^. However, as infection rates continue to increase there is a need for rapid antigen-based assays that can be performed at non-BSL3 point of care sites. Moreover, serological assays can promote the understanding of both T cell and B cell responses and lead to the isolation of neutralizing antibodies that can be later examined as therapeutics. Lastly, serological assays can highlight potential targets for vaccine candidates ^6-8^.

MPXV expresses approximately 25 structural proteins on the mature virion (MV), a form that is dominant during inter-host transmission, and additional 6 proteins on the enveloped virion (EV), a form with a second double membrane, that is dominant during intra-host transmission ^9,10^. Similar to other pox viruses, entry to host cells is mediated through interactions with glycosaminoglycans, and through fusion with plasma membrane at neutral pH (mostly EV), or through low pH-expedited endocytosis (mostly MV) ^11,12^. Studies conducted on the related Vaccinia virus (VACV) identified proteins important for viral attachment and entry ^11,12^ and several were found to be targeted by antibodies elicited in immunized mice, infected macaques and both infected and vaccinated humans ^13 14,15^. However, the main serological and B cell markers accompanying MPXV infection in humans are still not characterized.

To investigate the antibodies elicited following MPXV natural infection, we recruited a cohort of 11 convalescent donors 33-62 days post MPXV infection. All donors were diagnosed in Israel between May and June 2022 (Figure 1A). MPXV is highly similar to VACV which is the prototype of this viral family ^10^. Therefore, we first tested binding of heat-inactivated MPXV convalescent sera to heat-inactivated VACV-coated plates by enzyme-linked immunosorbent assay (ELISA) as previously described ^16^. As controls, we included serum samples of uninfected male donors below 40 years old (samples collected before the Monkeypox outbreak, n=5, named “uninfected <40yo”), as well as samples of uninfected male donors over 50 years of age (samples collected before the Monkeypox outbreak, n=6, named “uninfected >50yo”). The latter are expected to have been previously vaccinated with a Vaccinia-based vaccine, as until 1978 smallpox vaccination was a standard protocol in most countries, including Israel. IgG from heat-inactivated sera from all MPXV convalescent donors were able to bind VACV (Figure 1B). Sera from uninfected >50yo also recognized VACV, although to a lesser extent compared to MPXV convalescent samples. This agrees with previous reports demonstrating that vaccinated individuals exhibit high titers of antibodies even 50 years post vaccination ^17^. Sera from uninfected <40yo had minimal reactivity to the inactivated VACV.

**Figure 1:**
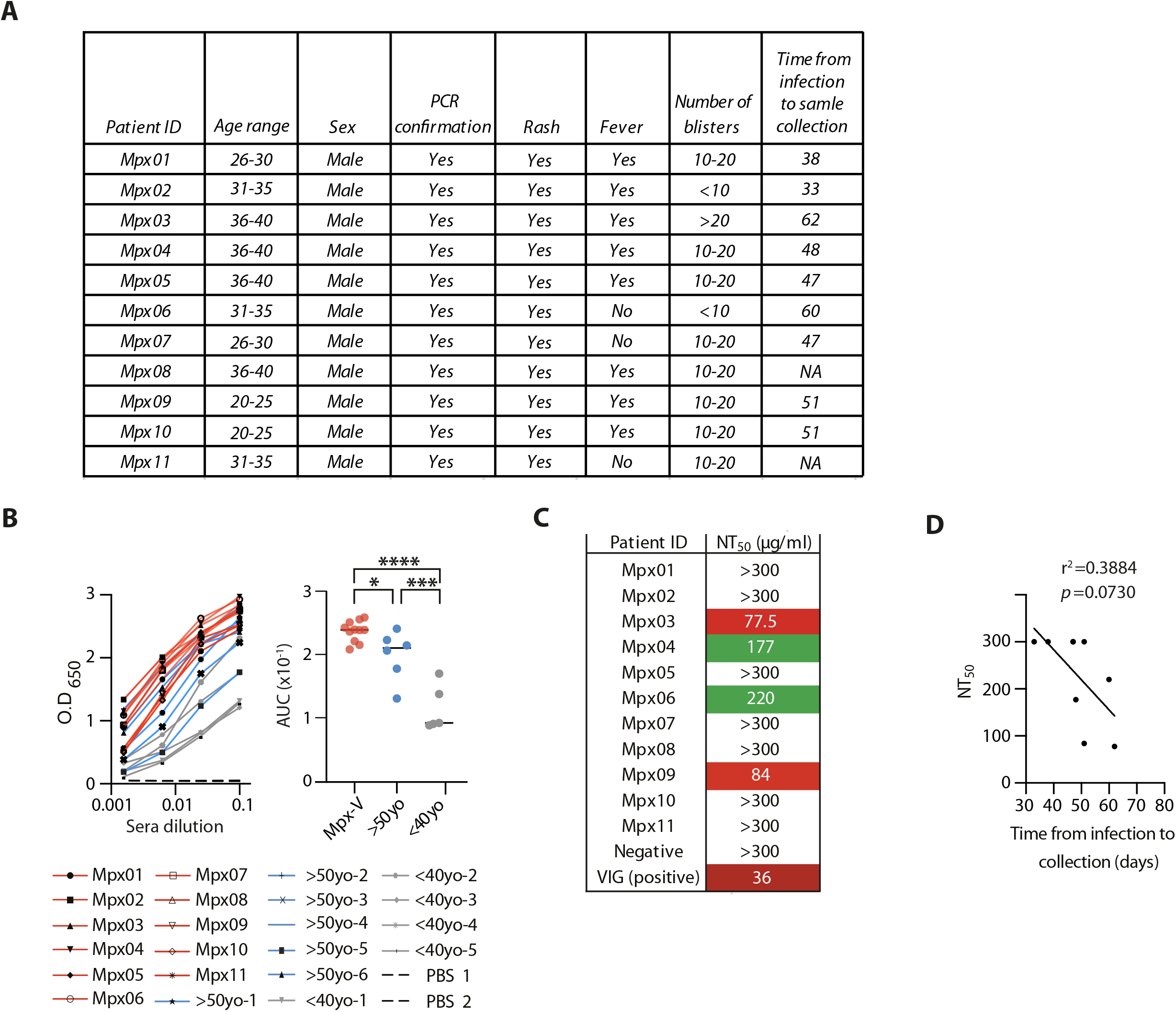
MPXV convalescent donors’ clinical data and serum IgG response to VACV virus. **(A)** Clinical data describing 11 MPXV convalescent donors recruited for this study.* Date of infection was provided by the patient upon clinical examination. NA – non applicable. **(B)** Sera responses by ELISA against inactivated VACV IHDJ strain ^16^. Left panel: binding curves of sera from MPXV convalescent donors (‘Mpx’, red lines), uninfected donors >50 years old (‘>50yo’, blue lines), and uninfected donors <40 years old (‘<40yo’, gray lines). PBS controls are in bold dashed lines. Plates were incubated with an HRP-conjugated anti-human IgG secondary antibody and following substrate addition, the OD was read at 650 nm. **(C)** Right panel: Area under the curve (AUC) values of the sera VACV IHDJ strain ELISA. Statistical analysis was carried out using one-way ANOVA test. **(D)** NT_50_ values in PRNA of VACV Lister strain by IgG purified from MPXV convalescent serum (see text for more details). ‘Negative’ represents purified IgG from serum of an uninfected donor <40 years old, ‘VIG’ represents Vaccinia Immune Globulin. PFU were counted after 72 hours and NT_50_ was determined. Color code is as follows: maroon, red, and green indicate high, mediocre and low neutralizing titers, respectively. White indicates no neutralization. **(E)** Correlation between time from infection to sample collection and donors NT_50_. Trend line represents simple linear regression. NT_50_ values >300 were plotted as 300. AUC, correlation, and r-squared values were calculated using GraphPad Prism software.

We next asked whether MPXV convalescent donors exhibit neutralizing antibodies. As a model we used Vaccinia Lister virus (VACV Lister) and determined neutralization of protein-A-purified IgG from MPXV convalescent sera in a plaque reduction neutralization assay (PRNA). Briefly, 50 plaque forming units (PFU) per well of VACV Lister were pre-incubated for 1 hour with purified IgG at a starting concentration of 200 μg/ml followed by 6 consecutive 2-fold dilutions and used to infect 5×10^5^ Vero cells. Purified IgG from an uninfected donor <40yo was used as negative control, while pooled purified IgG from VACV vaccinated donors was used as positive control (Vaccinia immune globulin, ‘VIG’ ^18^). The experiment was carried out in duplicates, and 72 hours post infection plaques were counted and NT_50_ was determined. Surprisingly, only 4 MPXV convalescent donors exhibited neutralizing activity (Figure 1C). NT_50_ values negatively correlated with the time from infection to sample collection, although this correlation was not statistically significant, possibly due to small sample size (Figure 1D). We conclude that although infection induces strong serological responses in ELISA, most of the recoverees do not have neutralizing antibodies against VACV Lister, 33-62 days post MPXV infection.

Next, we wished to map the targets for antibodies elicited following infection. Therefore, we generated soluble domains of four MPXV antigens that were previously described to be implicated in immune response against either MPXV or VACV ^13-15,19,20^. We produced two antigens from the EV form, A35R (95.03% homologous to VACV antigen A33R) amino acids 58-181, and A36R (97.02% homologous to VACV antigen A34R) amino acids 39-168, and two antigens from the MV form: M1R (98.4% homologous to VACV antigen L1R) amino acids 2-185 and H3L (93.52% homologous to VACV antigen H3) amino acids 2-277 (Figure 2A). The antigens were cloned into pcDNA3.1(-) expression vector, containing an 8xHistidine tag at their C-terminus followed by a 15-amino acid biotinylation sequence (Figure 2B). To validate that our truncated and tagged proteins acquire conformations similar to the structures of the native proteins as revealed by X-ray crystallography, we aligned the published PDB structures ^21-23^, to AlphaFold ^24^-predicted structures of the expressed proteins with tags. According to our analyses, the two conformations align well structurally (Figure 2C). The alignment for A36R was carried out with AlphaFold-predicted full-length protein, as no solved structure of this protein existed. The antigens were expressed in mammalian Expi293F cells and purified on Nickel beads, as previously described (Figure 2D) ^25^. The purified antigens were used to coat ELISA plates and reacted with sera IgG from the MPXV convalescent donors and both control groups. Sera IgG from all MPXV convalescent donors, bound A35R in a dose dependent manner (Figure 2E). While some uninfected >50yo donors reacted with A35R, overall, both uninfected donors’ groups exhibited significantly lower binding to this antigen. MPXV convalescent sera binding to H3L antigen was also detected. However, the signal was somewhat weaker when compared to A35R, with no statistical difference between MPXV convalescent donors and uninfected >50yo. We found no significant binding of MPXV convalescent sera to A36R or M1R (Figure 2E), except for uninfected >50yo group which exhibited statistically different titers to M1R compared to uninfected <40yo group. No sera IgG binding to any of the four antigens was detected amongst uninfected <40yo. Finally, no sera IgM activity against any of the produced antigens was detected in all groups (data not shown). Similar results were obtained with protein-A-purified IgG (Figure 2F). We conclude that both A35R and H3L are targets for antibodies elicited following infection, with the 124-amino acid truncation of A35R being able to distinguish between recent MPXV infection and past VACV vaccination.

**Figure 2:**
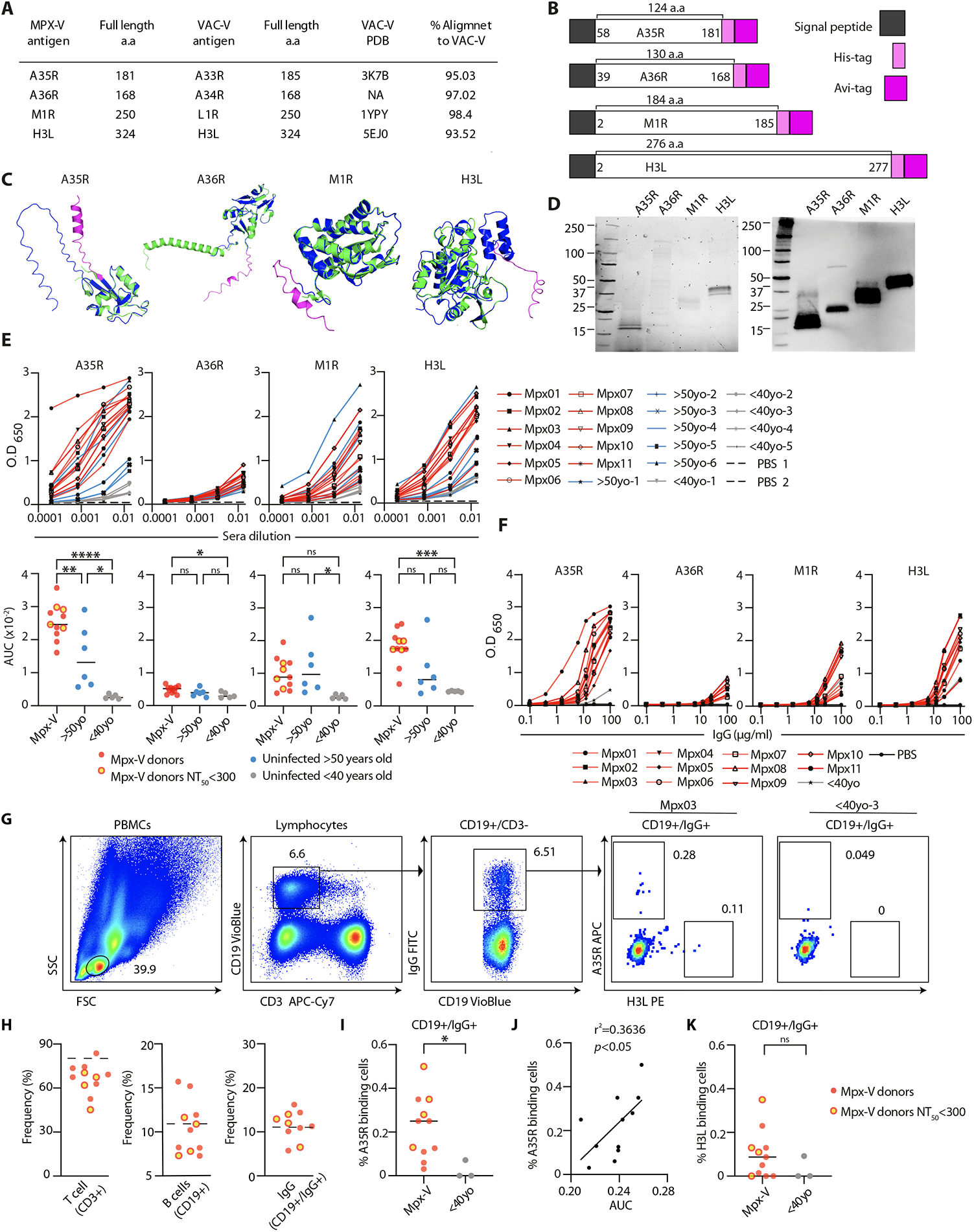
Antibodies and B cells from MPXV convalescent donors bind A35R and H3L antigens. **(A)** Table listing MPXV antigens A35R, A36R, M1R and H3L produced in this study. Published PDB IDs of VACV homologs are given. Alignment scores between MPXV and VACV antigens were calculated using the Clustal Omega web tool. NA – non applicable. **(B)** Construct design of the four produced antigens. Signal peptide is in dark gray, His-tag is in light pink and Avi-tag is in magenta. The amino acid section produced for each antigen is stated. **(C)** Cartoon representation of the 3-dimensional (3-D) structures of the four antigens A35R, A36R, M1R and H3L. In green are the published 3-D structures of the VACV homologs as described in Figure 2A. In blue are the AlphaFold predicted structures of the expressed antigens. His- and Avi-tags are presented in magenta. For A36R the alignment was carried out with AlphaFold predicted full-length protein. The two structures are superimposed one on top of the other. Images generated using PyMOL version 2.5.2. **(D)** Protein SDS-PAGE (left) and Western Blot (right) of the four antigens (500 ng) after purification. The antigens are indicated on top, the protein marker is on the left in each gel/blot, and protein sizes are indicated in kDa. **(E)** Top panel: Binding curves of sera from MPXV convalescent donors (‘Mpx’, red lines), of uninfected donors >50 years old (‘>50’, blue lines), and of uninfected donors <40 years old (‘<40’, gray lines) to four produced MPXV antigens (indicated on top). PBS controls are in black dashed lines. X axis is sera dilution, y axis is OD_650_. Plates were incubated with an HRP-conjugated anti-human IgG secondary antibody and following substrate addition, the OD was read at 650 nm. Bottom panel: Area under the curve (AUC) values of the serum ELISA. Yellow data points indicate MPXV recoverees who exhibited neutralizing activity in the plaque assay (Figure 1C). Statistical analysis was carried out using one-way ANOVA test. **(F)** Binding curves of purified IgG from MPXV convalescent donors (‘Mpx’, red lines), and one uninfected donor (‘<40’ in gray). PBS control is in a black line. X axis is IgG concentration, y axis is OD_650_. Plates were incubated with an HRP-conjugated anti-human IgG secondary antibody and following substrate addition, the OD was read at 650 nm. **(G)** Flow cytometry gating strategy for staining of A35R- and H3L-specific IgG+ B cells. Representative plots for Mpx03 donor are shown. A representative uninfected control is depicted on the right. **(H)** Left, middle and right panels: frequency of CD3+, CD19+ and IgG+ B cells (gated from CD19+ population), respectively, from PBMCs of MPXV convalescent donors. Dashed line represents cell frequency in an uninfected donor. Yellow data points indicate MPXV recoverees who exhibited neutralizing activity in the plaque assay (Figure 1C). **(I)** Frequency of A35R-specific IgG+/CD19+ cells in MPXV convalescent donors in (‘Mpx’, red), and uninfected donors (‘<40’, gray). Statistical analysis was carried out using unpaired T-test. **(J)** Correlation between AUC of VACV binding by ELISA (Figure 1B) and the frequencies of A35R-specific IgG+ B cells. Trend line represents simple linear regression. **(K)** Frequency of H3L-specific IgG+/CD19+ cells in MPXV convalescent donors in (‘Mpx’, red), and uninfected donors (‘<40’, gray). Statistical analysis was carried out using unpaired T-test. AUC, correlation and r-squared values were calculated using GraphPad Prism software.

Antibodies are produced by B cells following antigen encounter and activation ^26^. We therefore evaluated the ability of B cells collected from the 11 MPXV convalescent donors to specifically bind A35R and H3L antigens. For this purpose, 5-6 million PBMCs from whole blood were stained for CD3, CD19, IgG as described before ^25,27^, as well as with biotinylated, streptavidin fluorophore conjugated A35R and H3L (Figure 2G). MPXV donors exhibited frequencies of CD3+ cells ranging from 45%-83%, CD19+ cells ranging from 2%-10% and IgG B cells (gated from CD19+ population) ranging from 5%-16% (Figure 2H). We were able to record A35R-specific IgG+/CD19+ cells in all MPXV donors at frequencies ranging between 0.03% and 0.5% of the total IgG+/CD19+ cells (Figure 2I). Moreover, the frequencies of A35R-specific IgG+/CD19+ cells positively correlated with sera IgG VACV binding by ELISA (Figure 2J). Overall, lower levels of H3L-specific IgG+/CD19+ cells were detected in MPXV donors. Although the frequencies of H3L-specific IgG+/CD19+ cells were higher in MPXV donors than in controls, the difference was not statistically significant (Figure 2K). Three MPXV donors did not have any H3L-specific IgG+/CD19+ cells, indicating that B cell responses towards this antigen are less profound at the time samples were collected.

A35R, is one of the 6 proteins expressed on the EV form of the virus, which is believed to be responsible for cell-to-cell viral spread ^28^. While the EV form of poxviruses is considered more protected and more difficult to neutralize by antibodies ^29^, antibodies against EV proteins were found to mediate VACV neutralization after Vaccinia vaccination and MPXV infection ^13,30^ and removal of EV-directed antibodies abolished VIG neutralization ^31^. While most of the MPXV convalescent donors in our cohort did not exhibit neutralizing activity against VACV in a plaque assay 1-2 months after infection, all of them bound strongly to a 124-amino acid truncation of A35R antigen. Therefore, A35R was able to effectively distinguish MPXV infected donors. Additionally, we found a positive correlation between the frequency of anti-A35R B cells in peripheral blood and sera IgG binding to VACV by ELISA, indicating that at least some of the serological activity against VACV is contributed by A35R-specific B cells. These results suggests that the 124-amino acid truncation of the A35R antigen can be potentially used for antigen-based diagnosis of MPXV infection.

H3L antigen is expressed on the MV form, promoting binding to host cells and infectivity ^32^. It was identified as a target for both T cells and B cells ^14,20^ in vaccinated mice and humans, and anti-H3L antibodies were able to elicit protection from a lethal challenge in mice ^33^. In agreement with that, in our study, MPXV convalescent donors bound H3L, with no statistical difference between previously vaccinated and recently infected donors. Less MPXV donors exhibited H3L-specific IgG+ B cells than A35R-specific IgG+ B cells, with three exhibiting no H3L-specific IgG+ B cells at all. Amongst the MPXV donors who did not have any detectable H3L-specific IgG+ B cells was donor Mpx06 who showed neutralizing activity against the VACV Lister in plaque assay. This might suggest that other MPXV specific antibodies and T cells were elicited by infection and contributed to viral clearance.

In summary, in the present report we demonstrate that MPXV convalescent donors produce both antibodies and B cells against MPXV antigens A35R and H3L, and that a recombinant soluble truncated forms of A35R and H3L proteins can be used in ELISA as diagnostic tools for the detection of recently infected MPXV patients.

## Data Availability

All data produced in the present study are available upon reasonable request to the authors

## Ethics Statement

All MPXV convalescent donors between the ages of 23 and 39 provided a written informed consent prior to participating in this study. All donors tested positive in a PCR assay for MPXV infection collected from saliva, blood, anus, or blisters. A single sample of 170 mL of whole blood was collected 33-62 days post infection, 2-3 weeks after the donors were considered recovered. Tel Aviv University Institutional Review Board approved all studies involving patient enrollment, sample collection, and clinical follow-up. Tel Aviv University Institutional Review Board ethics approval has been granted (protocol number 0005243-1). Donors were followed by the Dermatology Division of Ichilov Tel Aviv Sourasky Medical Center (Helsinki approval number 0384-22-TLV). The uninfected >50 and uninfected <40 samples were collected from the Israeli Blood bank, Tel Aviv University Institutional Review Board approval protocol number 0004554-2. Ethics approval has been granted.

## Acknowledgments

We thank all the donors. We thank the members of the Freund Lab for fruitful discussions. We thank Dr Ohad Mazor from the Israeli Institute for Biological Research for his support. K.P.’s research is supported in part by a fellowship from the Edmond J. Safra Center for Bioinformatics at Tel-Aviv University. We thank Vice President of Research and Development of Tel Aviv University for funding. We thank Mr. Eli Gelman and Prof. Ariel Porat for their support.

## Conflict of interest

The authors declare no conflict of interest

## Notes

### Competing Interest Statement

The authors have declared no competing interest.

### Funding Statement

The study was funded by Tel Aviv University

### Author Declarations

Ethics Statement: All MPXV convalescent donors between the ages of 23 and 39 provided a written informed consent prior to participating in this study. All donors tested positive in a PCR assay for MPXV infection collected from saliva, blood, anus, or blisters. A single sample of 170 mL of whole blood was collected 33-62 days post infection, 2-3 weeks after the donors were considered recovered. Tel Aviv University Institutional Review Board approved all studies involving patient enrollment, sample collection, and clinical follow-up. Tel Aviv University Institutional Review Board ethics approval has been granted (protocol number 0005243-1). Donors were followed by the Dermatology Division of Ichilov Tel Aviv Sourasky Medical Center (Helsinki approval number 0384-22-TLV). The uninfected >50 and uninfected <40 samples were collected from the Israeli Blood bank, Tel Aviv University Institutional Review Board approval protocol number 0004554-2. Ethics approval has been granted.

### Summary of Updates

There has been a mistake in the order of the authors in the HTML. Additionally the middle initial of one of the authors was missing. The order of the authors is now correct. Look forward to see the revised version. All the best wishes, Natalia

